# Multi-organ structural and functional deficits in association with long COVID: a population-based case-control study

**DOI:** 10.64898/2026.02.12.26346170

**Authors:** Alba Fernández-Sanlés, Lucy J Goudswaard, Dylan M Williams, Betty Raman, Ellen J Thompson, Michele Orini, Siana Jones, Alexandra Jamieson, Lee Hamill Howes, Andrew Wong, Vedika Handa, Carole H. Sudre, Laura C Saunders, Nathan Cheetham, Alex Whitmarsh, Mary Ní Lochlainn, Jim Wild, Stephen M Smith, Stefan Piechnik, Stefan Neubauer, Claire J Steves, Nicholas J Timpson, Nish Chaturvedi, Alun D Hughes, the CONVALESCENCE, study collaborative

## Abstract

**Background:** Multi-system impacts of long COVID remain unknown.

We aimed to compare multi-system deficits between people with long COVID and controls.

**Methods:** We conducted a case-control study and recruited participants from two UK population-based cohorts: the Avon Longitudinal Study of Parents and Children (ALSPAC) and TwinsUK. Participants provided samples for SARS-CoV-2 serology between 2020 and 2021 and were asked about duration of COVID-19 symptoms between July and December 2021. Cases had long COVID (evidence of COVID-19 infection and persistent symptoms ≥4 weeks post infection); controls comprised 3 groups: acute COVID-19 only (symptoms reported for <4 weeks) and serological evidence of infection; self-reported long COVID-like symptoms but without wild-type SARS-CoV-2 virus antibodies; no symptoms or history of COVID-19 infection. People who were severely unwell or pregnant were excluded. Participants attended a clinic follow-up visit between 2021 and 2023 and underwent multi-system MRI, (cardiac, brain, lung, kidney), measurement of blood pressure and autonomic function, spirometry, renal function, exercise tests, strength and physical capability. Severity of deficit was then scored for each system as 0 (none) to 3 (severe). Primary outcome was a single composite multi-domain score summing each of nine domains: autonomic, brain, exercise capacity, heart, lungs, physical, renal, strength and vascular, with a maximum score of 27.

**Findings:** In total, 349 participants, 141 with long COVID (40%) and 208 (60%) controls were recruited. Overall deficit score in cases was 0.22 (95% CI -0.44,0.88) units greater than controls, adjusted for age, sex, ethnicity, cohort membership and relatedness. This estimate was little changed (0.32 (-0.34, 0.98)) when additionally adjusted for educational status, index of multiple deprivation, physical activity, smoking and co-morbidity. Restricting cases to those reporting symptoms including fatigue (n=46) increased the excess deficit score to 0.81 (-0.19,1.81) units in the minimally adjusted model. A difference was only observed in the vascular domain, largely attributable to elevated blood pressure, showing a 1.76 (1.04,2.97) multivariable adjusted odds ratio excess in cases, and 3.04 (1.36,6.80) when restricted to cases with fatigue.

**Interpretation:** There was no evidence of marked residual subclinical deficits in most systems in people with long-COVID, although there was evidence of persistent deficits in the vascular system, largely related to elevated blood pressure. Mechanisms unrelated to organ dysfunction may contribute to symptoms. Blood pressure measurement and control should be included in clinical follow-up.

**Funding:** Jointly funded by the National Institute for Health and Care Research and UK Research and Innovation (CONVALESCENCE, COV-LT-0009, MC_PC_20051).

**Research in context:** *Evidence before this study:* This study was established in 2021, at the height of the COVID-19 pandemic, when reports of persistent covid symptoms (Long COVID) in people ostensibly recovered from acute infection were recorded. We searched in PubMed from database inception to 28/02/24 for papers published in English using the terms ( "Post-Acute COVID-19 Syndrome"[MeSH] OR "long COVID"[tiab] OR "post-acute COVID"[tiab] OR "post-COVID syndrome"[tiab] OR PASC[tiab]) AND (multisystem[tiab] OR "multi-organ"[tiab] OR systemic[tiab] OR "multiple organ"[tiab]) AND (abnormalities[tiab] OR dysfunction[tiab]). Our search yielded 149 results. These included numerous studies and systematic reviews of both hard events and excess propensity to disease and subclinical disease in relation to Long COVID. These latter have been inconsistent, with findings ranging from little or no, to substantial abnormalities in various systems.

*Added value of this study:* Previous studies have often focussed on a single system and not taken a comprehensive deficit approach. Further, case definition has rarely been restricted to those reporting Long COVID, often recruiting from people hospitalised with COVID or from long COVID clinics. Control definitions have also been variable and may have introduced bias. We show minimal multi-system deficit in association with Long COVID, but in secondary analyses do observe a deficit in the vascular system, largely attributable to high blood pressure.

*Implications of all the available evidence:* People with Long COVID should be reassured that there is typically no persistent multi-system deficit as a consequence of their illness, however, they and their clinicians should pay attention to blood pressure which may warrant treatment.

## Introduction

While the global Coronavirus disease 2019 (COVID-19) pandemic due to SARS-CoV-2 has subsided, a substantial proportion of individuals continue to experience persistent and debilitating health effects consequent on infection during the pandemic^1,2^. This condition, variously referred to as long COVID or post-COVID syndrome, has conflicting definitions, but typically is characterised by persistence of multi-system symptoms for more than 4 to 12-weeks after initial infection.^3^ Here we adopt the term long COVID in line with current UK guidance.^4^ While estimates of the prevalence of long COVID and related conditions depend on its definition,^5^ it has been suggested that around 3% of the population of England and Scotland live with long COVID, with three-quarters reporting limitations of daily living, and a third reporting symptoms for at least 3-years.^6^

While COVID-19 infections now typically result in mild illness and full recovery, the risk of developing long COVID, though significantly reduced from a height of around 50-85% of hospitalised cases during the pandemic, remains unacceptably high in the post-pandemic vaccinated era at around 8-12%.^2^ The vast majority of COVID-19 cases, even during the pandemic, were not admitted to hospital. Long COVID is not restricted to those with severe infection, occurring in community-managed cases, and although risks are lower than in those hospitalised, the sheer number of not-hospitalised individuals means that most long COVID cases arise from community managed infections, a population that has hardly been studied.

While many individuals acquiring COVID-19 during the pandemic made a prompt recovery, in some recovery occurred many months or even years post infection; in others, recovery remains only partial or characterised by relapses. Even in those who lack symptoms or have subjectively recovered there may be evidence of persistent organ damage.^7^ The relationship between long COVID and organ deficits is unclear, and the extent to which persistent symptoms over months or years are associated with system deficits has not been adequately explored. This issue is further complicated by methodological issues such as the ascertainment of prior infection, definition and selection of long COVID sufferers and the choice of appropriate controls.

While risk factors for long COVID, including female sex, have been consistently identified,^8^ the syndrome itself remains poorly defined, and its underlying pathophysiological mechanisms, and multi-organ impacts are not fully understood. Previous studies of long COVID have largely recruited hospitalised cases of COVID-19,^9^ or those attending long COVID clinics,^10^ and variously report defects of different organ systems, including the lungs, cardiovascular system, brain, kidney and, in association, disordered cognitive and autonomic function at diagnosis of long COVID and up to 6 – 12 months later. Inconsistent findings may relate to small sample sizes, choosing controls from a different population from that which cases arose, variability in symptoms depending on severity and vaccination status,^11^ and inability to account properly for pre-existing co-morbidity.

We compared multi-organ measures of structure and function in a nested case-control study recruited from two population cohorts, Avon Longitudinal Study of Parents and Children (ALSPAC) and TwinsUK, hypothesising that people who had experienced long COVID would have a greater degree of multi-organ deficits than controls.

## Methods

### Study design

CONVALESCENCE (COroNaVirus post-Acute Long-term EffectS: Constructing an EvideNCE base) is a nested case-control study of 349 adults participating in two UK cohort studies, ALSPAC and TwinsUK. Protocol details are published,^12^ (https://rps.ucl.ac.uk/viewobject.html?cid=1&id=2393996) and briefly summarised here. ALSPAC is a multigenerational population-based cohort, that recruited pregnant women between 1991-1992, with repeat follow up both of this birth cohort and their parents.^13–15^ Participants were sent repeat COVID-19 questionnaires during the pandemic, which included a question on permission to re-contact. Home blood sampling kits were used for SARS-CoV-2 serology in April 2021. Those who had COVID-19 were asked about duration of symptoms between July and December 2021. TwinsUK is a countrywide twin registry established in 1992.^16^ Participants provided samples for SARS-CoV-2 serology between 2020 and 2021^17^ and were also asked for permission to re-contact. Invitations to participate in this deep phenotyping study were sent in batches to randomly selected individuals in each cohort who had agreed to re-contact, and for whom we had serological evidence of SARS-CoV-2 status. Those who were severely unwell or pregnant were excluded. Twins in TwinsUK were invited as a pair. A flowchart of participation is shown in Supplementary Figure S1.

Cases were defined as those who self-reported COVID-19 symptoms for ≥4 weeks, had serological evidence of infection (confirmed at clinic), and had no other known explanation for their symptoms. Three groups of controls were identified: *group 1* - acute COVID-19 only (symptoms reported for <4 weeks) and serological evidence of infection; *group 2* – long-term symptoms following COVID-19 (≥4 weeks in ALSPAC, ≥12 weeks in TwinsUK), but no serological confirmation of infection, *group 3* - no COVID-19 symptoms and no evidence of infection.

The overall CONVALESCENCE study was approved by South Central – Oxford A Research Ethics Committee (21/SC/0235). In addition, each cohort underwent individual ethical review, ALSPAC from the ALSPAC Ethics and Law Committee, ref B3666, and TwinsUK from HRA/NHS Research Ethics Committees, TwinsUK Biobank 19/NW/0187, TwinsUK EC04/015 or Healthy Ageing Twin Study 07/H0802/84.

### Data collection

Participants attended the Bloomsbury Centre for Clinical Phenotyping (BCCP) in central London between 2021 and 2023. Participants gave written informed consent to the study at the beginning of the clinic visit. Questionnaire data were collected using REDCap^18^ on symptoms (see Supplementary Table S1), sociodemographic factors, physical activity (using the International Physical Activity Questionnaire – Short Form (IPAQ-SF), co-morbidity (including cardiovascular disease, diabetes (type 1 or type 2), hypertension, dyslipidaemia, asthma, and chronic obstructive pulmonary disease; all either defined as diagnosed by a doctor, and/or on medication), COVID-19 vaccination status, and time since COVID-19 infection. Local area deprivation score was assigned based on postcode of residence. Participant sex was self-reported.

Height, weight and body fat on bioimpedance were measured to a standardised protocol. Magnetic resonance imaging (MRI) of lungs, heart, brain, cardiovascular system and kidneys was performed, and post-processed aligned to the C-MORE protocol.^19^, which investigated post-hospitalised individuals with COVID-19.

Spirometry and MRI measured lung function. Brachial and central blood pressure and heart rate were measured in the seated, lying and standing positions. A 12-lead electrocardiogram (ECG) plus 5-minute recording for heart rate variability was captured. Exercise capacity and cardiopulmonary fitness tests were performed on those without contra-indications. Handgrip assessed strength, while chair rises and ability to stand on one leg for several seconds assessed physical status. Serum creatinine was used to calculate the estimated glomerular filtration rate.^20^

### Deficit scores based on deep phenotypes

Deficits for each domain were pre-specified (Supplementary table S2). Where possible, deficit-informing variables derived from the deep phenotypes were defined by a knowledge-informed approach (based on established thresholds widely used in clinical practice or reported in the literature). Otherwise, we used a data-driven approach, defining thresholds from the measurements in the control subsample. Severity of deficit was then scored for each domain as follows, none [0], mild [1], moderate [2] and severe [3]. A single composite multi-domain score was calculated as our primary outcome, summing each of 9 domains: autonomic, brain, exercise capacity, heart, lungs, physical, renal, strength and vascular; thus a maximum score of 27 could be obtained.

### Sample size and power

The original planned sample size of 800 participants had to be revised downwards as the pandemic progressed, since few if any participants escaped infection. The revised sample size combined all control groups. Assuming a 1:1 balance of cases versus controls, using a 27-unit continuous deficit scoring system, the revised sample of 350 participants would allow detection of a standardised difference in multi-organ score of ≥0.3, equivalent to a one-unit difference *between* cases and controls with 80% power and 5% significance. Based on the scoring system we used, an average difference of one unit in deficit severity could correspond to a one step difference in a single domain (e.g. none versus mild, or mild versus moderate), or to a much smaller average difference distributed across multiple domains. A one-unit difference was therefore considered the smallest deficit that would be deemed clinically important.

### Statistical analyses

Analyses were conducted using R 3.6.2 and Stata 18 within the secure UCL trusted research environment (Data Safe Haven).

Participant characteristics were compared between cases and the combined controls on complete data for that variable, using a Student’s *t*-test for continuous variables, and a Chi-squared test for categorical variables. Cells reporting data on fewer than 5 people are reported as <5. In primary analyses, missing data on phenotypes were imputed with multiple imputation by chained equations (50 imputed datasets with the predictive mean matching method and five iterations). Associations between case/control status and overall multi-domain system deficit score were modelled using linear mixed models in multiple imputed data. Domain specific binary scores (normal (0) versus abnormal (1-3)) were calculated using logistic regression, again in multiple imputed data. Three models were constructed for each outcome; Model 1: adjusted for age, sex, ethnicity, membership of either ALSPAC or TwinsUK cohorts, and relatedness, Model 2: Model 1 plus educational status, index of multiple deprivation, physical activity and smoking status, Model 3: Model 2 plus comorbidity. Relatedness (twin pairs in TwinsUK, and mother/partners/offspring clustering in ALSPAC) was accounted for in models with a random effect intercept for family membership in linear mixed models and robust standard errors in logistic regression models.

Five secondary analyses were performed. First, we compared median deficit score for cases, and each of the three original control groups. Second, a complete data analysis (i.e. without multiple imputation). Third, a comparison of outcomes in cases restricted to those with any persistent symptoms at the time of clinic and fourth, cases restricted to those reporting fatigue at clinic — relative to all controls in each instance (with multiple imputation of data). Fifth, we compared all cases to controls excluding the control subgroup reporting long COVID-like symptoms without evidence of SARS-CoV-2 infection (again with multiple imputation). Sex-stratified analyses were not performed due to the preponderance of women in the study. Inference was based on size of effect, precision of estimates and p values. No adjustment was made for multiple testing.

## Results

### Sample characteristics

In total, 349 people, 141 participants with long COVID (40%) and 208 (60%) controls were recruited (Table 1). On average cases and controls were in their early 50s, mostly of European ethnicity, and more likely to be women. While education levels were similar, cases were more likely to reside in disadvantaged neighbourhoods. Cases reported fewer co-morbidities. Most cases and controls had been vaccinated by the time of clinic attendance. At clinic, time since COVID-19 infection was shorter in cases (median 14.1 weeks) than the controls who reported prior infection (19.9 weeks), and 69% of cases versus 24% of controls reported any symptoms (table 1 and figure 1), with fatigue being the most frequent symptom, reported by 46/141 cases (33%). Missingness rates by phenotypes overall varied from 0% in the vascular domain, to 40% for the autonomic domain (Supplementary table S3).

**Figure 1.**
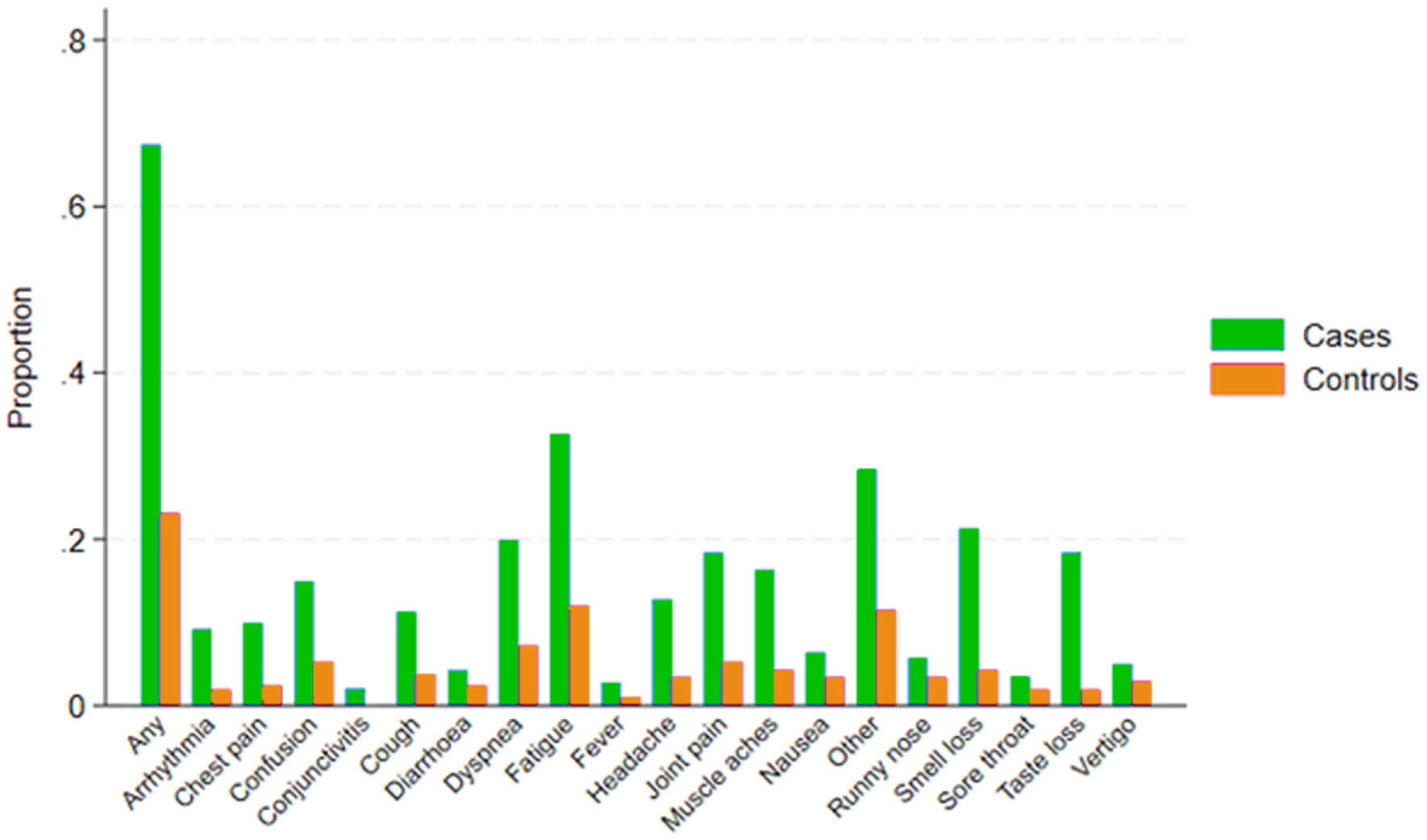
Self-reported symptoms associated with long COVID between cases and controls.

**Table 1.**
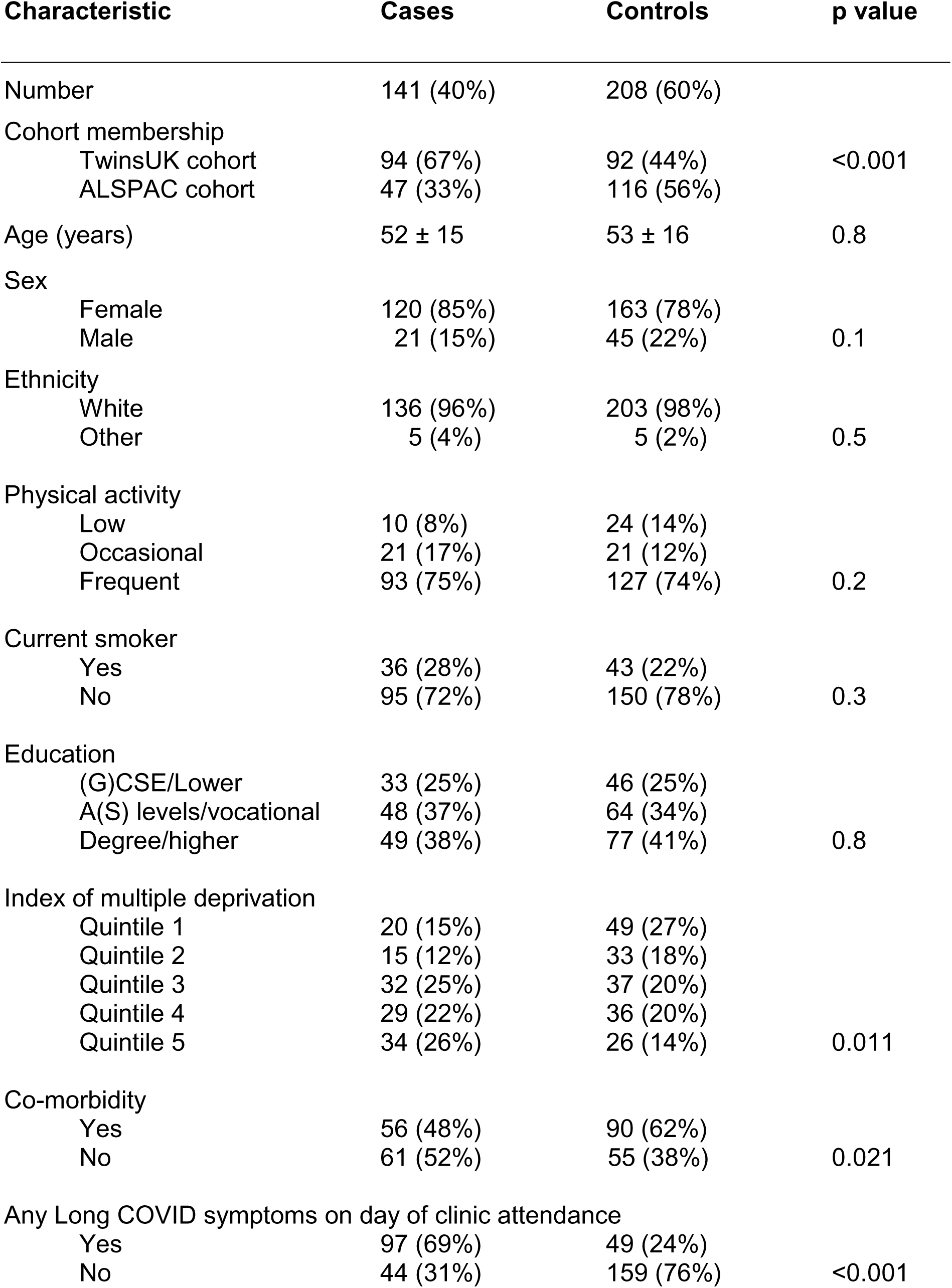
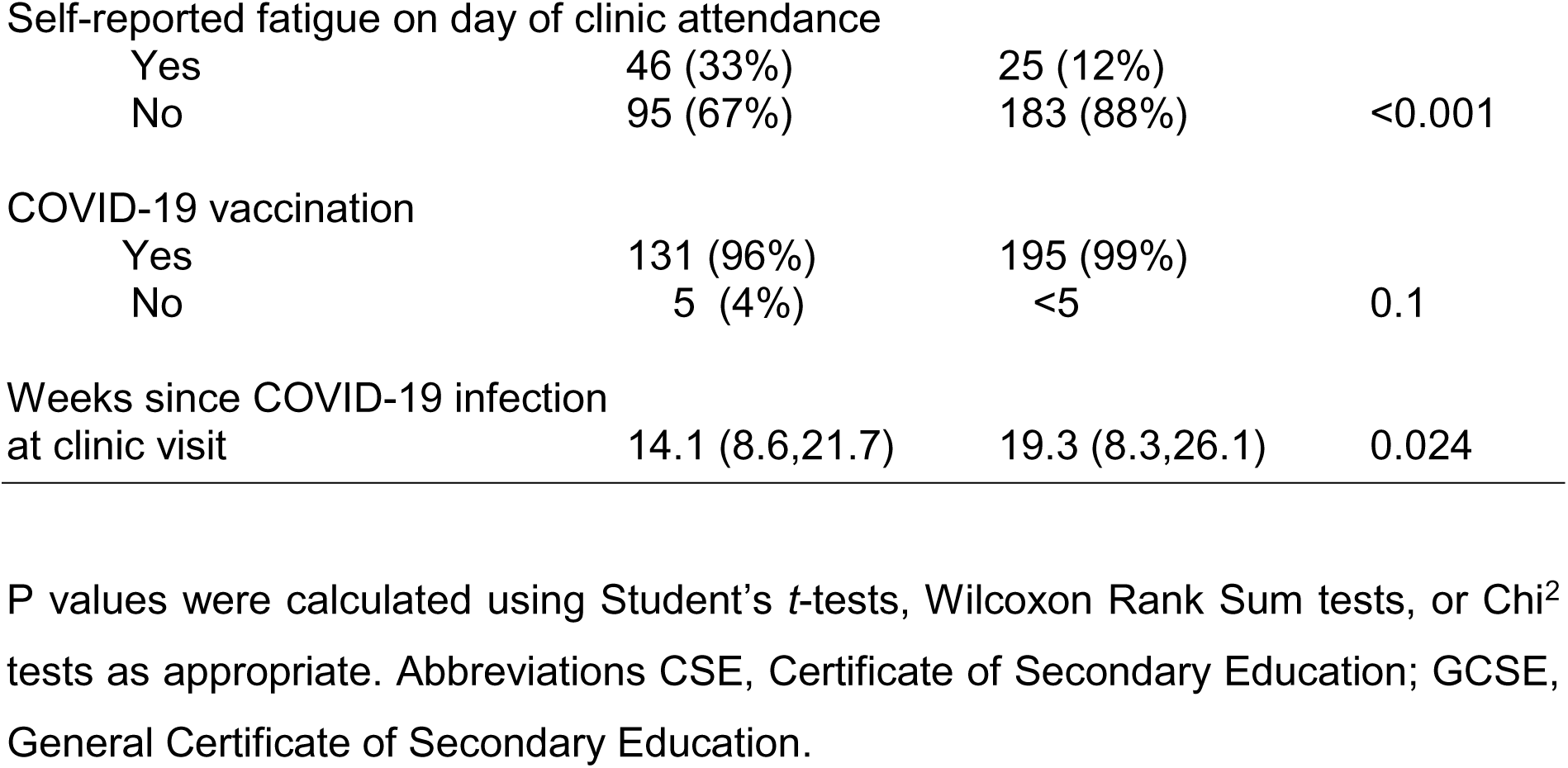
Participant characteristics by case control status - means ± SD, median (25^th^,75^th^ centiles), or n (%)

### Association between long COVID and deficit scores

The composite deficit score in the minimally adjusted model was higher by 0.22 units (95% CI _-_0.44, 0.88) in long COVID cases compared to controls (figure 2A) but confidence limits were wide. Adjustment for socioeconomic, health behaviour characteristics and co-morbidity barely altered this estimate (0.32 (-0.34, 0.98) units greater in cases than controls). The most striking domain specific difference between cases and controls was in the vascular domain, with multivariable-adjusted odds ratio of 1.75 (1.04, 2.97) (figure 2B). This was attributable to an increased odds of hypertension (odds ratio of 2.05 (0.89, 4.71) after full adjustment in the long COVID group.

**Figure 2.**
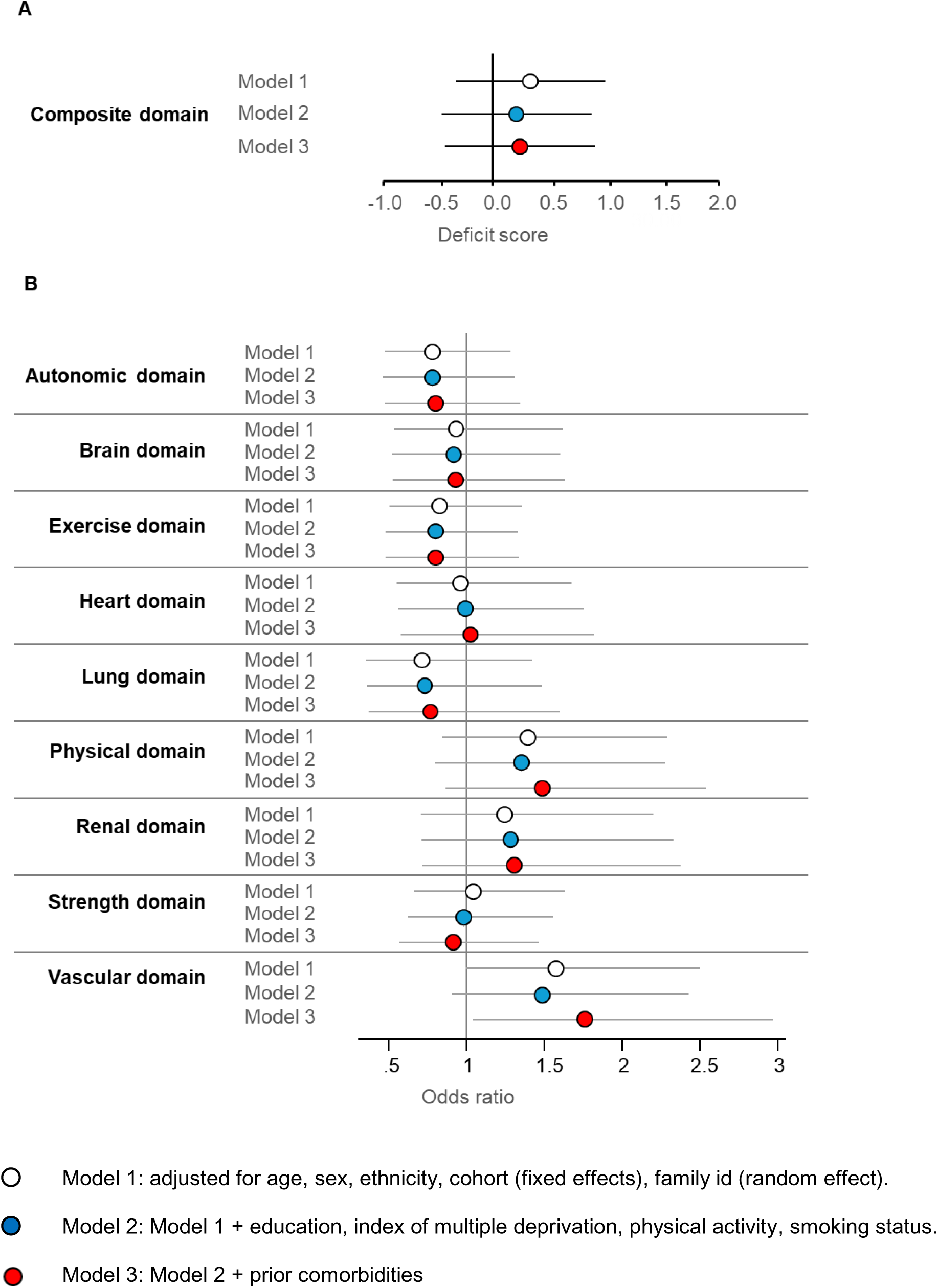
Panel A) Mean overall deficit difference and panel B) Odds ratio by domain comparing cases and controls (with 95% confidence limits).

### Secondary analyses

Median (inter-quartile range) deficit score did not differ by case/control status, nor when controls were sub grouped by presence of symptoms and by infection status; being 5 (2,7) for cases, 3 (1,4) for those recovered from COVID-19 within 4 weeks, 4 (3,6) for those with long COVID symptoms but no evidence of COVID-19 infection, and 4 (2,7) for those without symptoms or evidence of COVID-19 infection (Supplementary table S3).

When we restricted the analysis to those with complete data on the measures of target organ deficit (Supplementary table S4), the composite deficit score was similar to that observed in the multiply imputed dataset (0.25 units (-0.82,1.33) greater in cases than controls in the minimally adjusted model, increasing to 0.85 (-0.24,1.93) with multivariable adjustment, with wide confidence limits.

Restricting the case definition to those reporting any long COVID symptoms on the day of clinic attendance (97/141) and using the multiply imputed dataset (Supplementary table S5) made only a marginal difference to the overall score difference, being 0.37 (-0.39,1.06) units greater in cases than controls in the minimally adjusted model; additional multivariable adjustment made little difference to this estimate (0.41 (-0.31. 1.13) units). As in the primary analysis, the vascular domain was the most different between cases and controls, with an odds ratio of 1.97 (1.10, 3.52) after full adjustment. Further restricting cases to those reporting symptoms that included fatigue at clinic (46/141) additionally increased the overall score difference to 0.69 (-0.27, 1.66) units in the fully adjusted model (Supplementary table S6). Again the vascular domain was the most affected, with a fully adjusted odds ratio of 3.04 (1.36, 6.80).

Restricting the definition of controls to exclude all those having reported long COVID-like symptoms historically, but with no evidence of COVID-19 infection (44/208) (Supplementary table S7) barely changed the overall score deficit difference between cases and controls, being 0.32 (-0.37,1.01) in the minimally adjusted model, and 0.45 (-0.24,1.14) in the full model. The vascular domain was most affected, with a multivariable odds ratio of 1.94 (1.10,3.42).

## Discussion

Excess major system events, specifically cardiometabolic, neurological and respiratory, and compromised exercise capacity have been observed up to one year following COVID-19 infection.^21–27^ This, and the observation of ongoing multi-system symptoms in those with long COVID potentially imply important underlying subclinical deficits that could persist even if symptoms improve. We found little evidence of marked composite multi-system deficits based on target organ structure and function in people with long COVID compared to controls. Our primary analysis suggests an average difference between cases and controls of less than a third of a unit on a 27-unit deficit scale across multiple systems. A difference of this size roughly corresponds to a 3–4% increase in the probability that an individual is rated one severity category worse in any domain. However, this small average difference masks considerable between system heterogeneity; with a close to twofold higher odds of vascular deficit in cases.

Our findings appear somewhat discordant with the reported greater risks of events and functional compromise across multiple systems in people surviving symptomatic COVID-19 infection. There are several possible explanations. First, long COVID symptoms and subclinical disease may not be closely linked. Previous report of excess events or compromised function post COVID-19 have limitations, notably that they have not usually distinguished between all those infected and those with long COVID. A similar limitation applies to studies of subclinical disease, where there is greater heterogeneity of findings, ranging from no specific abnormality to severely compromised function^26,28–31^ and no studies, including ours, have pre-infection deep phenotyping measures. Systematic reviews also highlight biases, including residual confounding, differences in definition of target system deficit phenotype, severity of infection, vaccination status and imprecise case and control definition (for those studies that include controls), as reasons for heterogeneity and inability to perform a meta-analysis. Despite only small multidomain differences, we did observe an excess risk of vascular disease, largely attributable to elevated blood pressure. It is notable that two previous meta-analyses of new onset hypertension post COVID-19 infection both reported an increased risk (hazard ratio and odds ratio respectively) of 1.70,^27,32^ though again it should be noted that these studies included a mix of people post COVID-19, not all of whom declared long COVID.

Most previous studies of subclinical disease have focussed on hospitalised patients, those attending long COVID and other clinics, or those with persistent system-specific symptoms.^9,33^ A cross-system comparison of people who had not been hospitalised in association with COVID-19 infection found little in the way of significant target organ damage, and notably nothing on MRI imaging.^34^ Similarly, the RECOVER-Adult cohort found no difference in clinical blood and urine biomarkers in people with post-acute sequelae of COVID-19 compared to those without.^35^ In general, more severe disease (i.e. requiring hospitalisation), is associated with evidence of persistent subclinical target organ damage, and, given the persistent symptom reporting in long COVID, it could be argued that a similar excess of target organ damage would be anticipated. However, there are important differences between long COVID and severe COVID-19 infection. Women, and young to middle aged adults are more likely to suffer long COVID, but may also be more resilient to subclinical disease, whereas severe infection and clinical disease is more frequent in men and older people.^8^

Second, our case group was a mix of people who were and were not experiencing symptoms on the day of assessment. Persistence of symptoms is a widespread and complex issue following illness.^36^ In our sample, 2/3 of long COVID cases reported symptoms with 1/3 reporting fatigue, the most frequent long COVID symptom. This does not necessarily imply that these individuals had fully recovered since long COVID is frequently characterised by a relapsing-resolving pattern.^37^ Restricting cases to those attending clinic and reporting any symptoms slightly increased the overall difference between cases and controls from 0.32 to 0.41 units (fully adjusted models) but with reduced precision. Restricting analysis to those with symptoms including persistent fatigue further increased the difference to 0.69 but with wide confidence intervals that were compatible with no difference.

A third explanation relates to control rather than case selection. We recruited a mix of control groups, including people who reported recovering from acute COVID-19 with no ongoing symptoms, and people reporting symptoms but with no serological evidence of prior COVID-19 infection, as well as non-infected and asymptomatic people. If fully recovered infection, and/or symptoms without evidence of infection are also associated with subclinical disease, this could serve to reduce differences between cases and controls. However, selecting individuals without previous infection or symptoms could also create a bias by using a ‘super-healthy’ comparator group. Nevertheless, in our analysis, the overall deficit score was similar in all control groups, and similar to that observed in cases. Excluding the symptomatic control group did not materially alter the overall difference in deficit score between cases and controls.

Fourthly we had to reduce the original sample size as the pandemic progressed, and recruitment of truly uninfected controls became impossible. Our revised power calculation enabled detection of at least a small to moderate difference in deficit score between cases and controls. The overall difference in deficit score between cases and controls for the primary analysis was far smaller than this, at around a 1/3 of a unit. While this was compatible with no difference we also cannot exclude a small difference.

Most previous studies have focussed on single organs, or mechanistically connected systems, such as cardiopulmonary measures. In CONVALESCENCE, we studied multiple systems concurrently, reflecting the diverse potential impacts of infection. We endeavoured where possible to categorise deficits using clinically meaningful cut-points. For example, a 10 mmHg systolic blood pressure difference has been associated with a 1.2 fold excess risk of cardiovascular events^38^, with a similar excess risk observed for a category change in estimated glomerular filtration rate^39^. The lowest quintile of heart rate variability is associated with a >1.5 fold excess risk of death^40^, while inability to perform a static balance test is associated with ∼1.1 fold excess risk of mortality^41^. Thus a one unit difference in our 27 unit scale is likely to be clinically important, so our observation of an average 0.69 units of deficit in people with symptoms that included fatigue is small but may not be negligible. Further, a 1.7 to 3-fold excess of vascular deficit has potentially important clinical implications. Nevertheless, establishing clinically meaningful cut-points for many of our measures was challenging. In some instances we had to resort to using our control group to establish cut-points to define deficits. If controls were more similar to cases (in terms of either experiencing symptoms, or seemingly having recovered relatively quickly from COVID-19 infection), this would serve to minimise any difference between cases and controls. We should also note that hypertension was included as an outcome in the vascular domain, as people with treated hypertension may well have resting pressures within the normal range. It is difficult to precisely date the onset of hypertension, and we included a history of hypertension as a co-morbidity to account for this. Strikingly, controls had a greater burden of co-morbidity, thus if anything this will have resulted in an underestimate of any difference between cases and controls in the vascular domain.

Key strengths of our study include studying multiple systems simultaneously using a multimodal approach (imaging and non-invasive physiology measures), recruitment from established cohorts, independent validation of COVID-19 infection, as clear as possible a definition of long COVID case status, and multiple secondary analyses recognising the limitations of defining both a case and control.

We conclude that, 3-4 months post COVID-19 infection, there is little evidence of marked overall system deficit in people who have experienced long COVID. This might imply that other mechanisms apart from organ dysfunction contribute to persistent symptoms in long COVID. There was some evidence of excess elevated blood pressure in cases which may be of clinical importance and should therefore be attended to in people with long COVID.

## Data Availability

Due to the sensitive nature of the data collected for this study, data cannot be made publicly available, but requests to access the dataset from qualified researchers trained in human subject confidentiality protocols may be sent to at the Unit for Lifelong Health and Ageing at UCL.

## Contributors

Alba Fernández-Sanlés - data curation, formal analysis, writing – original draft

Lucy J Goudswaard - formal analysis, writing– review & editing

Dylan M Williams - formal analysis, validation, writing– review & editing

Betty Raman - conceptualisation, formal analysis, validation, writing– review & editing

Ellen J Thompson - formal analysis, writing– review & editing

Michele Orini - writing– review & editing

Siana Jones - investigation, writing– review & editing

Alexandra Jamieson - investigation, writing– review & editing

Lee Hamill Howes - investigation, project administration, resources, writing– review & editing

Andrew Wong - investigation, project administration, resources, writing– review & editing

Vedika Handa - formal analysis, writing– review & editing

Carole H. Sudre - formal analysis, writing– review & editing Laura C Saunders - formal analysis, writing– review & editing

Nathan Cheetham - formal analysis, writing– review & editing Alex Whitmarsh - formal analysis, writing– review & editing

Mary Ní Lochlainn - writing– review & editing

Jim Wild- supervision, writing– review & editing

Stephen M Smith - supervision, writing– review & editing

Stefan Piechnik - supervision, writing– review & editing

Stefan Neubauer - writing– review & editing

Claire J Steves - conceptualisation, funding acquisition, supervision, writing– review & editing

Nicholas J Timpson - conceptualisation, funding acquisition, supervision, writing– review & editing

Nish Chaturvedi - conceptualisation, formal analysis, funding acquisition, project administration, supervision, validation, writing– review & editing

Alun D Hughes - conceptualisation, formal analysis, funding acquisition, project administration, supervision, validation, writing– review & editing

Alba Fernández-Sanlés and Dylan M Williams accessed and verified the underlying data.

Alex Whitmarsh could not be contacted so his forms could not be completed, but all other authors vouch for his contribution.

## Acknowledgements

We are extremely grateful to all the people who took part in this study, and to the past and present members of the research team who collected and managed the data.

