## Supplementary figures and tables for "Multi-organ structural and functional deficits in association with long COVID: a population-based case-control study"

**Supplementary table S1 - Symptoms elicited at clinic (in alphabetical order).**

Arrhythmias/palpitations

Chest pain

Confusion/difficulties concentrating

Conjunctivitis/sore eye

Cough

Fatigue

Headache

Joint pain

Loss of smell

Loss of taste

Low grade fever

Muscle aches

Nausea/lack of appetite

Other

Runny nose

Shortness of Breath

Sore throat

Vertigo

### Supplementary table S2. Items comprising the nine health domains

| DOMAINS | | SELECTED VARIABLE(S) | | | | DERIVED VARIABLES | | | DOMAIN-SPECIFIC SCORE |
|---|---|---|---|---|---|---|---|---|---|
| **Health domain** | **Sub-domain** | **Trait** | **Variable/s** | **Measure/s** | **Number of times measured** | **Derived Metric/s [item coding]** | **Criteria used to define thresholds;<br>thresholds (if relevant)** | **Reference Paper<br>(if relevant)** | **0 – no deficit, 1 – mild deficits; 2 – moderate deficits; 3 – severe deficits** |
| **1) Autonomic function** | **Postural blood pressure (BP)** | Orthostatic hypotension | Change in peripheral BP from lying to standing after 1min and 3min | Lying systolic BP (SBP) / diastolic BP (DBP)<br>Standing SBP/DBP | 3 (repeated)<br>2 (2 timepoints) | Orthostatic hypotension (Change in peripheral BP from lying [average of 2nd and 3rd measures] to standing after 1min or 3min)<br>[0/1] | Guidelines;<br>Orthostatic hypotension: decrease in SBP by ≥20 mmHg, or decrease in DBP by ≥10 mmHg, within 3min after standing<br>(change ≤ -20 SBP / -10 DBP at 1 min or at 3 min coded as 1) | https://academic.oup.com/bmb/article/115/1/123/260117 | 0: No deficits<br>1: deficit in 1 metric<br>2: deficits in 2 metrics<br>3: deficits in at least 3 metrics |
| | **Heart rate** | Sinus tachycardia | Resting heart rate while seated (HR) | Seated HR | 3 | Sinus tachycardia [ 0/1] | Guidelines; Mean of three seated HR >100 = 1 | 2015 ACC/AHA/HRS Guideline for the Management of Adult Patients With Supraventricular Tachycardia \| Circulation | |
| | | Dysfunctional heart rate variability (HRV) | Time and frequency metrics on HR profiles | Root mean square of successive differences between normal-to-normal intervals (RMSSD) | 1 | HRV abnormality [0/1] | Literature; low RMSSD <25 ms | https://pubmed.ncbi.nlm.nih.gov/20663071/;<br>ps://pubmed.ncbi.nlm.nih.gov/20663071/;<br>https://www.frontiersin.org/articles/10.3389/fpubh.2017.00258/full | |
| | | Dysfunctional heart rate recovery (HRR) | Change from maximum HR to 1min after stopping the CPET test | Change from maximum HR to 1min after stopping the CPET test | 1 | HRR abnormality [0/1] | Guidelines;<br>HRR ≤25 bpm = 1 | https://www.nejm.org/doi/full/10.1056/nejmoa043012 | |
| | | Postural tachycardia | Change in HR from lying to standing after 1min and 3min | Lying HR<br>Standing HR | 3 (repeated)<br>2 (2 timepoints) | POTS<br>[ 0/1] | Guidelines;<br>Postural tachycardia: Increase in HR by ≥30 bpm, within 3min after standing<br>(change ≥30 at 1 min or at 3 min coded as 1) | https://www.ncbi.nlm.nih.gov/pmc/articles/PMC9012474/ | |
| **2) Vascular** | **Resting Blood pressure** | Seating peripheral BP | Average peripheral BP | Seating peripheral SBP/DBP | 3 | Hypertension groups (mean of 2nd and 3rd BP measures) [0/1/2/3] | Guidelines; ≥140 SBP / 90 DBP = 3; 130-140/80-90 = 2; 120-130 & <80 = 1; <120 = 0 | https://www.heart.org/en/health-topics/high-blood-pressure/understanding-blood-pressure-readings;<br>https://www.escardio.org/Journals/E-Journal-of-Cardiology-Practice/Volume-17/definition-of-hypertension-and-pressure-goals-during-treatment-esc-esh-guidelin | 0: history=0 & hypertension ranges=normal<br>1: history=0 & hypertension ranges=mild<br>2: history=0 & hypertension ranges=moderate<br>3: history=1 or hypertension ranges=severe |
| | | Hypertension<br>Hypertensive medication | Self-reported hypertension<br>Self-reported hypertensive medication | Hypertension<br>Use of antihypertensive | 1<br>1 | History of hypertension (history or medication) [ 0/1] | | | |
| **3) Exercise Response** | **Cardiopulmonary exercise test (CPET)** | % Predicted peak VO2 | Predicted VO2 max achieved using age, sex, weight and height | Predicted peak VO2 | 1 | Predicted peak VO2<br>[coded 0/1/2/3] | Guidelines;<br>0: >84% ; 1: >70%, ≤84%; 2: >50%, ≤70%; 3: ≤50% or meeting exclusion criteria for CPET | https://respiratory-research.biomedcentral.com/articles/10.1186/s12931-021-01895-6 | Scored as per predicted peak V02 categories |
| **4) Muscle strength** | **Bioimpedance** | Low percentage muscle mass (PMM) | | TABC_PMM | | Low muscle mass<br>[0/1] | PMM <40% = 1 | https://www.ncbi.nlm.nih.gov/pmc/articles/PMC8350199/ | 0: no deficits; 1: deficit in one item; 3; deficit in both item (NB: omits second category) |
| | **Hand-grip strength** | Hand-grip strength | | Hand-grip strength | 3 | Hand-grip strength based on maximum test value of three<br>[0/1] | Literature; Sex- and age-dependent cut-offs for low grip strength (<10th percentile) = 1 | https://journals.plos.org/plosone/article?id=10.1371/journal.pone.0113637 | |
| **5) Physical function** | **Sit-to-stand test** | FTSTS or 5xSTS (five times sit to stand test)<br>TTSTS or 10xSTS (ten times sit to stand test) | Time to complete up to 10 chair rises (also available time for 5) | chair rises | 1 | Chair rises<br>[0/1] | Literature; age-dependent cut-offs (age<70, 5 rises in ≥11.4 seconds =1; age 70-79, 5 rises in ≥12.6 seconds =1; age≥80, 5 rises in ≥14.8 seconds = 1) | https://pubmed.ncbi.nlm.nih.gov/17037663/<br>https://journals.lww.com/nsca-jscr/Fulltext/2011/11000/Test_Retest_Reliability_of_the_Five_Repetition.36.aspx | 0: no deficits; 1: deficit in one item; 3; deficit in both item (NB: omits second category) |
| | **Balance exercises** | Flamingo balance exercises | Flamingo balance exercises: time for balance with one leg raised with eyes open and closed | Flamingo balance exercises | eyes open: 2 per leg<br>eyes closed: 1 per leg | Balance<br>[0/1] | Literature; age-dependent cut-offs; performance with open eyes<15 seconds or performance with closed eyes <5 seconds = 1 | https://pubmed.ncbi.nlm.nih.gov/19839175/ | |
| **6) Brain** | **Brain MRI** | Incidental findings | | Incidental findings | 1 | Incidental findings [0/1] | | | Domain score = highest score between WBV and WMHV items; anyone with incidental findings coded 3 irrespective of WBV and |
| | | Whole brain volume (WBV) | | WBV | | Whole brain volume<br>[0/1/2/3] | Data-driven; Based on distributions of this variable in control group 3 (no report of COVID-19 from recruitment) without comorbidities, thresholds at <10th centile [1], <5th centile [2] or <1st centile [3] | https://pubmed.ncbi.nlm.nih.gov/19839175/l29 | |

| | | | | | | | | | |
|---|---|---|---|---|---|---|---|---|---|
| | | Cerebrovascular lesions | | Total white matter hyperintensities | | White matter hyperintensities [0/1/2/3] | Data-driven; Based on distributions of this variable in control group 3 (no report of COVID-19 from recruitment) without comorbidities, thresholds at >90th centile [1], >95th centile [2] or >99th centile [3] | [illegible] | [illegible] WMHV scores |
| **7) Heart** | **Cardiac MRI** | Incidental findings | | Incidental findings | 1 | Incidental findings [0/1/3] | Anyone with advisory CV-related note = 1; anyone with urgent CV-related note = 3 | NA | 0: no deficits or incidental findings; 1: advisory incidental findings only; 2: 1 deficit across items, but no advisory findings; 3: 2 or more deficits across items, or urgent incidental finding |
| | | T1 (longitudinal magnetization relaxation time constant) | | T1 Global | | T1 Global outlier [0/1] | Data-driven; 1 = participants >2 standard deviations from mean based on distributions of these variables in control group 3 (no report of COVID-19 from recruitment) without comorbidities | | |
| | | T2 ((transverse magnetization relaxation time constant) - global and middle | | T2 Global | | T2 Global outlier [0/1] | | | |
| | | Late gadolinium enhancement (LGE) | | Late gadolinium enhancement (LGE) | | LGE [0/1] | Any enhancement classified as abnormal | | |
| | | Left & right ventricular ejection fractions (R/LVEF) | | Left & right ventricular ejection fractions (R/LVEF) | | EF [0/1] | LVEF<50 = 1, RVEF<35 = 1 | https://www.sciencedirect.com/science/article/pii/S1097664723010529?via%3Dihub | |
| | **Resting ECG** | ECG abnormalities | | ECG abnormalities | 1 | Incidental findings [0/1] | Any participant with an advisory note from clinical review = 1 | NA | |
| **8) Pulmonary** | **Spirometry** | Forced Expiratory Volume in the 1st second (FEV1)<br>Forced Vital Capacity (FVC) | Externally derived z-scores for deviations from age- and sex-specific reference values from the GLI (https://gli-calculator.ersnet.org/index.html) | Z-FEV1 against external reference ranges<br>Z-FVC against external reference ranges<br>Z-FEV1/FVC ratio against external reference ranges | 1 | FEV1 [0/1]<br>FVC [0/1] | For each of z-FEV1, z-FVC, zFEV1/FVC: 0= measure ≥-1.645; 1= measure <-1.625 & measure ≥-2; 2 = measure <-2 & measure ≥-3; 3 = measure <-3 | https://pubmed.ncbi.nlm.nih.gov/22743675/ | Domain score = highest score from zFEV1, zFVC and zFEV1/FVC categories |
| | **Lung MRI** | Lung parenchymal abnormality | Qualitative axial thoracic HASTE images | four categories; up to 25%, 26-50%, 51%-75% and 76-100% | 1 | 0-4 | | https://doi.org/10.1016/s2213-2600(23)00262-x | |
| **9) Renal** | **Renal MRI** | Renal corticomedullary differentiation | Difference in the visualization of cortex and medulla; abnormality can indicate nephropathy | Renal corticomedullary differentiation | 1 | CMD in the left and right kidneys (mean) | Data-driven; Based on distributions of this variable in control group 3 (no report of COVID-19 from recruitment) without comorbidities, thresholds at <10th centile [1], <5th centile [2] or <1st centile [3] | NA | |
| | **Renal function** | eGFR | | eGFR | 1 | | 0 = eGFR ≥60 ; 1 = eGFR ≥45 & eGFR<60, 2 = eGFR ≥30 & eGFR<45, 3 = eGFR<30 | https://www.sciencedirect.com/journal/kidney-international-supplements/vol/3/issue/1 | Domain score = highest score between CMD and eGFR categories |

**Supplementary table S3 – Overall median deficit score, and domain specific presence or absence of deficit by case and control subgroup status**

| Outcome measure | | N (non-missing) | Case-control group | | | |
|---|---|---|---|---|---|---|
| | | | Cases<br>N = 141 | Recovered COVID-19 control_1<br>N = 75 | Long COVID like symptoms no COVID-19 infection control_2<br>N = 44 | No symptoms, no COVID-19 infection control_3<br>N = 89 |
| Overall deficit score, median (25th, 75th quantiles) * | | 142 | 5 (2, 7) | 3 (1, 4) | 4 (3, 6) | 4 (2, 7) |
| Autonomic domain, N (%) | | 210 | | | | |
| | no deficits | | 44 (57.9%) | 15 (65.2%) | 33 (57.9%) | 32 (59.3%) |
| | any deficits | | 32 (42.1%) | 8 (34.8%) | 24 (42.1%) | 22 (40.7%) |
| Brain domain, N (%) | | 306 | | | | |
| | no deficits | | 85 (66.9%) | 26 (68.4%) | 49 (62.8%) | 45 (71.4%) |
| | any deficits | | 42 (33.1%) | 12 (31.6%) | 29 (37.2%) | 18 (28.6%) |
| Exercise domain, N (%) | | 282 | | | | |
| | no deficits | | 65 (60.7%) | 17 (47.2%) | 40 (51.9%) | 43 (69.4%) |
| | any deficits | | 42 (39.3%) | 19 (52.8%) | 37 (48.1%) | 19 (30.6%) |
| Heart domain, N (%) | | 278 | | | | |
| | no deficits | | 86 (76.8%) | 26 (72.2%) | 53 (76.8%) | 47 (77.0%) |
| | any deficits | | 26 (23.2%) | 10 (27.8%) | 16 (23.2%) | 14 (23.0%) |
| Lung domain, N (%) | | 270 | | | | |
| | no deficits | | 96 (89.7%) | 27 (84.4%) | 60 (87.0%) | 51 (82.3%) |
| | any deficits | | 11 (10.3%) | 5 (15.6%) | 9 (13.0%) | 11 (17.7%) |

| | | | | | |
|---|---|---|---|---|---|
| Physical function domain, N (%) | 328 | | | | |
| no deficits | | 72 (55.0%) | 23 (54.8%) | 50 (60.2%) | 42 (58.3%) |
| any deficits | | 59 (45.0%) | 19 (45.2%) | 33 (39.8%) | 30 (41.7%) |
| Renal domain, N (%) | 287 | | | | |
| no deficits | | 92 (79.3%) | 29 (82.9%) | 65 (87.8%) | 50 (80.6%) |
| any deficits | | 24 (20.7%) | 19 (45.2%) | 9 (12.2%) | 12 (19.4%) |
| Strength domain, N (%) | 333 | | | | |
| no deficits | | 89 (67.9%) | 29 (70.7%) | 60 (69.0%) | 56 (75.7%) |
| any deficits | | 42 (32.1%) | 12 (29.3%) | 27 (31.0%) | 18 (24.3%) |
| Vascular domain, N (%) | 349 | | | | |
| no deficits | | 64 (45.4%) | 19 (43.2%) | 52 (58.4%) | 34 (45.3%) |
| any deficits | | 77 (54.6%) | 25 (56.8%) | 37 (41.6%) | 41 (54.7%) |

* from a total of 27 potential deficits

**Supplementary table S4 – Difference in overall deficit score and domain specific odds ratios between cases and controls – excluding those with missing values (complete case analysis, N=82)**

| | Model 1 beta, (95% CI) | Model 2 beta, (95% CI) | Model 3 beta, (95% CI) |
|---|---|---|---|
| **Difference in overall deficit score** | 0.25 (-0.82,1.33) | 0.24 (-0.87,1.35) | 0.85 (-0.24,1.93) |
| **Domain specific odds ratio** | | | |
| **Autonomic** | 1.31 (0.62,2.77) | 1.28 (0.55,2.95) | 1.33 (0.57,3.12) |
| **Brain** | 1.44 (0.73,2.85) | 1.42 (0.71,2.84) | 1.43 (0.71,2.89) |
| **Exercise** | 0.60 (0.31,1.15) | 0.53 (0.26,1.08) | 0.52 (0.25,1.08) |
| **Heart** | 1.09 (0.52,2.30) | 1.07 (0.51,2.22) | 1.15 (0.55,2.41) |
| **Lung** | 0.43 (0.17,1.10) | 0.46 (0.18,1.18) | 0.51 (0.19,1.37) |
| **Physical** | 0.76 (0.38,1.51) | 0.84 (0.39,1.80) | 0.96 (0.44,2.09) |
| **Renal** | 1.27 (0.58,2.82) | 1.34 (0.58,3.13) | 1.38 (0.60,3.21) |
| **Strength** | 0.97 (0.55,1.72) | 0.93 (0.52,1.66) | 0.87 (0.48,1.58) |
| **Vascular** | 1.50 (0.85,2.66) | 1.47 (0.79,2.73) | 1.75 (0.92,3.33) |

**Supplementary table S5 – Difference in overall deficit score and domain specific odds ratios between cases and controls – restricting cases to those reporting any symptoms at clinic (97/141)**

| | Model 1 beta, (95% CI) | Model 2 beta, (95% CI) | Model 3 beta, (95% CI) |
|---|---|---|---|
| **Difference in overall deficit score** | 0.37 (-0.39,1.06) | 0.31 (-0.40,1.02) | 0.41 (-0.31,1.13) |
| **Domain specific odds ratios** | | | |
| **Autonomic** | 0.81 (0.46,1.41) | 0.79 (0.44,1.40) | 0.80 (0.45, 1.44) |
| **Brain** | 0.98 (0.54,1.77) | 0.95 (0.52,1.73) | 0.96 (0.52,1.75) |
| **Exercise** | 0.95 (0.54,1.77) | 0.92 (0.53,1.63) | 0.92 (0.52, 1.75) |
| **Heart** | 0.96 (0.51,1.84) | 0.98 (0.51,1.88) | 1.01 (0.52,1.95) |
| **Lung** | 0.50 (0.20,1.26) | 0.51 (0.20,1.34) | 0.52 (0.20,1.40) |
| **Physical** | 1.34 (0.76,2.34) | 1.30 (0.73,2.33) | 1.38 (0.77,2.49) |
| **Renal** | 1.23 (0.66,2.28) | 1.26 (0.66,2.41) | 1.27 (0.66,2.42) |
| **Strength** | 0.92 (0.54,1.56) | 0.89 (0.52,1.52) | 0.83 (0.48,1.44) |
| **Vascular** | 1.80 (1.06,3.04) | 1.69 (0.97,2.95) | 1.97 (1.10,3.52) |

**Supplementary table S6 – Difference in overall deficit score and domain specific odds ratios between cases and controls – restricting cases to those reporting symptoms at least one of which was fatigue at clinic (46/141)**

| | Model 1 beta, (95% CI) | Model 2 beta, (95% CI) | Model 3 beta, (95% CI) |
|---|---|---|---|
| **Difference in overall deficit score** | 0.75 (-0.23,1.73) | 0.65 (-0.32,1.62) | 0.69 (-0.27,1.66) |
| **Domain specific odds ratios** | | | |
| **Autonomic** | 0.80 (0.39,1.64) | 0.75 (0.35,1.60) | 0.75 (0.36,1.60) |
| **Brain** | 0.87 (0.39,1.94) | 0.81 (0.36,1.84) | 0.81 (0.35,1.86) |
| **Exercise** | 1.22 (0.61,2.45) | 1.19 (0.57,2.46) | 1.18 (0.57, 2.47) |
| **Heart** | 1.01 (0.43,2.38) | 1.02 (0.44,2.40) | 1.02 (0.43,2.41) |
| **Lung** | 0.56 (0.14,2.29) | 0.61 (0.15,2.57) | 0.61 (0.15,2.60) |
| **Physical** | 1.27 (0.60,2.67) | 1.18 (0.54,2.54) | 1.17 (0.54,2.51) |
| **Renal** | 1.12 (0.51,2.43) | 1.20 (0.52,2.76) | 1.21 (0.52,2.77) |
| **Strength** | 0.84 (0.40,1.78) | 0.77 (0.35,1.68) | 0.75 (0.33,1.70) |
| **Vascular** | 3.02 (1.46,6.26) | 2.83 (1.27,6.28) | 3.04 (1.36,6.80) |

**Supplementary table S7 – Difference in overall deficit score and domain specific odds ratios between cases and controls – excluding 44 controls who had symptoms at clinic**

| | Model 1 (beta, 95% CI) | Model 2 (beta, 95% CI) | Model 3 (beta, 95% CI) |
|---|---|---|---|
| **Difference in overall deficit score** | 0.32 (-0.37,1.01) | 0.28 (-0.40,0.96) | 0.45 (-0.24,1.14) |
| **Domain specific odds ratios** | | | |
| **Autonomic** | 0.80 (0.47,1.35) | 0.79 (0.46,1.36) | 0.82 (0.47,1.41) |
| **Brain** | 0.92 (0.53,1.62) | 0.91 (0.51,1.61) | 0.92 (0.52,1.65) |
| **Exercise** | 0.92 (0.55,1.55) | 0.89 (0.53,1.51) | 0.89 (0.52,1.53) |
| **Heart** | 1.00 (0.55,1.81) | 1.04 (0.57,1.90) | 1.07 (0.58,1.97) |
| **Lung** | 0.76 (0.37,1.55) | 0.76 (0.37,1.57) | 0.82 (0.39,1.74) |
| **Physical** | 1.39 (0.82,2.35) | 1.36 (0.77,2.38) | 1.54 (0.86,2.79) |
| **Renal** | 1.20 (0.66,2.18) | 1.27 (0.67,2.40) | 1.28 (0.67,2.45) |
| **Strength** | 1.08 (0.68,1.70) | 1.02 (0.64,1.63) | 0.93 (0.57,1.51) |
| **Vascular** | 1.66 (1.02,2.69) | 1.58 (0.94,2.67) | 1.94 (1.10,3.42) |

**Supplementary Figure S1 – Flow chart of participation.**

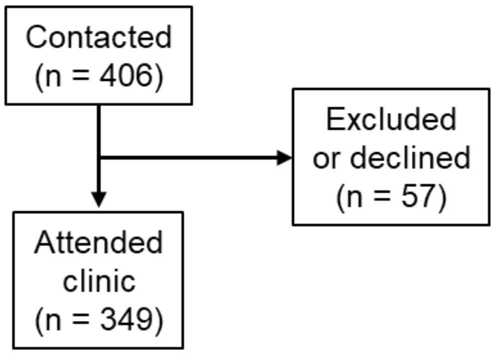
